# Biological processes linking soft drink consumption with site-specific cancer risk within the Global Cancer Update Programme (CUP Global)

**DOI:** 10.64898/2026.07.13.26356051

**Authors:** Emma Fontvieille, Nahid Ahmadi, Yahya Mahamat-Saleh, Nadine Hashem, Béatrice Lauby-Secretan, Marc J. Gunter, Fred K. Tabung, Suzanne D. Turner, Dieuwertje E. Kok, Lee Jones, Zdenko Herceg, Richard Simpson, Doris S. M. Chan, Konstantinos K. Tsilidis, Ahmad Jayedi, Christelle Clary, Helen Croker, Panagiota Mitrou, Elio Riboli, Stephen Hursting, Sarah Lewis, Laure Dossus

**Affiliations:** Nutrition and Metabolism Branch, International Agency for Research on Cancer, Lyon, France; Evidence Synthesis and Classification Branch, International Agency for Research on Cancer, Lyon, France; Department of Epidemiology and Biostatistics, School of Public Health, Imperial College London, London, UK; Division of Medical Oncology, Department of Internal Medicine, The Ohio State University College of Medicine, and Comprehensive Cancer Center, Columbus, USA; Division of Cellular and Molecular Pathology, Department of Pathology, University of Cambridge, Cambridge, UK; Faculty of Medicine, Masaryk University, Brno, Czech Republic; Division of Human Nutrition and Health, Wageningen University & Research, Wageningen, The Netherlands; Department of Population Sciences, Beckman Research Institute of the City of Hope, Duarte, USA; Epigenomics and Mechanisms Branch, International Agency for Research on Cancer, Lyon, France; School of Nutritional Sciences and Wellness, College of Agriculture and Life Sciences, University of Arizona, Tucson, USA; Department of Hygiene and Epidemiology, School of Medicine, University of Ioannina, Ioannina, Greece; World Cancer Research Fund International, London, UK; Nutrition Research Institute, University of North Carolina at Chapel Hill, Kannapolis, North Carolina, USA; Department of Population Health Sciences, Bristol Medical School, University of Bristol, Bristol, UK

**Keywords:** Review, soft drinks, biological pathways, mechanisms, insulin, glucose, pancreatic cancer, endometrial cancer, colorectal cancer, colon cancer, rectal cancer, ovarian cancer, kidney cancer, breast cancer, CUP Global, WCRF

## Abstract

This review evaluates the biological pathways linking soft drink consumption with the risk of several cancers within the framework of the Global Cancer Update Programme (CUP Global). Soft drink consumption has been associated with increased risk of multiple cancers, and glucose or insulin dysregulation has been proposed as a potential underlying mechanism.

We applied a three-stage framework. In the first stage, we identified insulin sensitivity as the key biological process potentially linking soft drink consumption (sugar-sweetened or artificially sweetened) to cancer risk, with glucose-related and insulin-related biomarkers as potential intermediate phenotypes, using a combination of expert knowledge and a web-based text mining tool. In the second stage, we conducted targeted PubMed searches to identify studies examining associations between consumption of soft drinks and these intermediate phenotypes (IPs) and between these IPs and the risk of several cancers in adult humans. In the third stage, the evidence was evaluated by the Expert Committee on Cancer Mechanisms (MEC), who assessed the strength of the evidence for these associations. The MEC concluded that there was weak evidence supporting a role of glucose or insulin-related processes as a potential mechanistic pathway linking the consumption of sugar-sweetened or artificially sweetened beverages to the risk of various cancers evaluated.

## Introduction

Nearly 20 million new cancer cases and 10 million cancer-related deaths occurred worldwide in 2022.(1) An estimated 30–50% of cancers are linked to modifiable lifestyle and environmental factors, indicating substantial prevention potential.(2) Among dietary factors, soft drinks consumption (including sodas, fruit drinks and other beverages with added sugars), the leading source of added sugars in the diet (3), has increased globally in recent decades.(4)

High intake of soft drinks increases the risk of adverse health outcomes, including overweight, obesity, and type 2 diabetes.(3,5,6) Increasing evidence also links sugar-sweetened beverages (SSB) to higher risks of several cancers(7–9) while evidence for artificially sweetened beverages (ASB), often used as alternatives to SSB, remains limited.(10)

Beyond their effect on body weight, both SSB and ASB may have an impact on cancer risk by inducing metabolic disturbances, in particular dysregulation of insulin and glucose levels.(11–18) but no comprehensive literature review has summarised mechanistic evidence from human studies linking soft drink consumption with cancer risk.

The Global Cancer Update Programme (CUP Global) by WCRF International (WCRFI) evaluates how modifiable risk factors such as diet, nutrition, physical activity, and body weight, affect cancer risk and survival. In CUP Global, the evaluation of plausible biological mechanisms is a key component of the review process that supports drawing causal inferences and determining the overall strength of the epidemiological evidence.(19)

As part of the CUP Global evaluation of soft drink consumption and cancer incidence (Jayedi et al., unpublished), we conducted a review of the biological processes underlying the association between soft drink consumption and the incidence of pancreatic, colorectal, endometrial, breast, kidney, or ovarian cancer.

## Methods

The methodology followed a framework developed within CUP Global (19) that aims at identifying and assessing the strength of evidence that the intermediate phenotypes (IPs) operate as a plausible biological mechanism between the exposure (soft drinks, as defined in supplementary material 1) and cancer site of interest.

The methodology comprised three stages: (i) identification of the IPs of interest; (ii) systematic reviews of human studies assessing associations between soft drink intake and the selected IPs, as well as between these IPs and the incidence of the cancers of interest; and (iii) evaluation of the strength of evidence for each biological process by the CUP Global Expert Committee on Cancer Mechanisms (MEC).

### Stage 1

A literature search was conducted using PubMed (see Supplementary Material 2) to identify studies reporting associations between soft drink consumption, potential IPs, and cancers of the pancreas, colorectum and endometrium. The retrieved articles were uploaded into a web-based tool called Text Mining for Mechanisms Prioritisation (TeMMPo), developed to quantify the volume of literature investigating a link between exposure and outcomes to a predefined list of IPs.(20) The results from this analysis were then presented to the MEC Chair and Deputy-Chair, who selected the IPs of interest for systematic review in stage 2, based on the biological plausibility and the amount of available literature.

### Stage 2

PubMed searches were conducted to identify studies investigating two sets of associations: first, between soft drink consumption and the IPs selected in stage one; and second, between those IPs and cancers of the pancreas, colorectum and endometrium. The list of cancer sites was then extended, based on epidemiological evidence of associations between soft drink consumption and cancer risk (Jayedi et al. unpublished), to kidney, ovarian and breast cancers. Detailed search strategies are provided in Supplementary Material 3. Titles and abstracts retrieved at this stage were screened independently by two reviewers, and full-text articles meeting the eligibility criteria were selected for data extraction. Inclusion and exclusion criteria are detailed in Supplementary Material 4. The eligibility criteria differed in the two sets of studies because IPs are short-term, modifiable outcomes suitable for randomized controlled trials (RCTs), whereas cancer is a long-latency outcome that is predominantly studied using observational designs (cohort and Mendelian Randomization (MR) studies) because of ethics and feasibility.

Following data extraction (supplementary material 5), we conducted a narrative synthesis of the evidence, summarising the findings of the associations between soft drink consumption and IPs, and between these IPs and site-specific cancer risk. In addition, forest plots were constructed including studies comparing the highest versus the lowest exposure categories to facilitate visualization of the associations between IPs and cancer risk; studies using other exposure measures (e.g., continuous) were described in the footnotes.

### Stage 3

Based on the narrative synthesis from Stages 1 and 2, the MEC evaluated the strength of evidence that the selected biological process operates as a mechanism between soft drink consumption and cancer outcomes. A decision-support tool was used to structure the judgement following a three-step process: (i) grading the strength of the exposure–IP and IP–outcome associations; (ii) integrating these gradings to assess overall biological plausibility; and (iii) issuing a conclusion on the strength of evidence (see Supplementary Material 6).(19)

## Results

Stage 1 results from the automated screening of literature on soft drinks and cancers of the colorectum (either specified as colon or rectal, or colorectal), pancreas, and endometrium are presented in Supplementary Figure 1. Glucose- or insulin-related intermediate phenotypes were among the top three IPs for the amount of available literature linking soft drinks to cancer risk with a larger number of abstracts identified linking IPs to cancer risk (>1000) but a sufficient number of abstracts linking soft drinks to IPs (>100). Based on these results as well as on expert opinion on biological plausibility, it was decided to focus on glucose- and insulin-related intermediate phenotypes for Stage 2 (definitions of IPs listed in supplementary material 1).

After screening and reviewing 3,375 studies on soft drink consumption and IPs, 10 prospective studies and 14 RCTs met the eligibility criteria (Supplementary Table 1 for prospective studies and Supplementary Table 2 for RCTs). The sample size of the RCTs ranged between 50 and 240 and the duration of the interventions ranged between 4 weeks and 12 months. Most prospective studies include >1000 participants, add a duration between the exposure assessment and IP measurement >4 years and presented results adjusted for adiposity.

A total of 21 studies (13 RCTs and 8 prospective cohort studies) assessed associations between soft drink consumption and glucose-related biomarkers.

Higher fasting blood glucose (FBG) levels in ASBs consumers compared to water consumers were observed in only one RCT out of four (21–24). Three RCTs from one research team showed that substitution of habitual ASBs consumption with water for 6 to 18 months among participants with overweight or obesity induced a reduction in 2-h postprandial (2hpp) glucose, while just one of them reported a reduction in glycated hemoglobin (HbA1c).(21–23)

Two prospective cohort studies reported positive associations between ASBs consumption and FBG or HbA1c (25,26) while another study found no evidence of an association.(27)

FBG levels were similar in RCTs comparing SSBs with water (two trials)(21,28) or with ASBs (two trials)(21,29). No effect was observed on FBG levels when replacing sucrose with high-fructose corn syrup in SSB beverages (one trial).(30) One RCT evaluating substitution of water for SSBs reported no strong evidence for an effect on HbA1c levels.(28)

Among prospective cohorts, two studies reported positive associations between carbonated SSBs consumption and FBG levels.(31,32). In contrast, two other studies found no evidence of an association between SSBs and FBG.(27,33) For carbonated soft drinks (unspecified sweeteners), one study observed a positive association with FBG (34), whereas another study reported no association (35).

A total of 14 studies (10 RCTs and 4 prospective cohort studies) examined the association between soft drink consumption and fasting insulin, C-peptide, and other insulin-related biomarkers.

For RCTs, there was no strong evidence that consuming SSBs affected insulin levels or indices of HOMA-insulin sensitivity and HOMA-B compared with unsweetened beverages (1 RCT).(21) Similarly, no differences were observed when comparing ASBs with SSBs (2 RCTs)(21,29) or when replacing sucrose with high-fructose corn syrup (1 RCT)(30). In prospective cohort studies, positive associations were found between HOMA-IR and the consumption of carbonated soft drinks (1 study)(34) or SSBs (2 studies)(34,36). One study did not observe strong evidence of an association between the intake of carbonated soft drinks and insulin levels.(34)

A reduction in insulin levels or HOMA-IR was observed when ASBs were replaced with water, among participants with overweight and obesity, in three RCTs from the same research team.(22–24) In two other RCTs, there was no strong evidence that consuming ASBs affected insulin levels or indices of insulin resistance compared with unsweetened beverages.(21,37)

In prospective cohort studies, no evidence of associations was reported between insulin-related IPs and ASBs consumption.(26,34,36,38).

Regarding studies on IPs and pancreatic cancer incidence, out of 4,943 abstracts, 24 prospective cohort studies and 3 MR studies met the inclusion criteria (Supplementary Table 3; Figures 1 and 2). Two additional relevant studies were identified by reference list review.

Six studies from Korea (22,775 cases) and one from China (512 cases) investigated the association between glucose levels and pancreatic cancer risk; all reported estimates above one when comparing the highest vs. the lowest categories (RR ranging from 1.08 and 2.52 with the confidence intervals not including the null for a large majority of them).(39–45) A Korean cohort study also found that hyperglycaemia measured repeatedly during follow-up was associated with an increased pancreatic cancer risk.(46) Three European prospective studies (1,590 cases) similarly reported effect estimates ranging from 1.24 to 2.39.(47–49) In contrast, two small cohort studies from Japan (10 cases) and the Netherlands (9 cases) did not show any evidence of an association.(50,51)

For HbA1c, six studies from Europe and the USA (1,836 cases overall) reported effect estimates for pancreatic cancer ranging from 1.02 to 2.42, although one study included the null in its confidence interval.(52–57) Another study suggested that HbA1c levels may increase several years before diagnosis.(58)

An MR study of participants of European ancestry reported an odds ratio (OR) of 1.24 (95% CI: 0.95–1.61) per 1-standard-deviation (SD) increase in genetically proxied fasting glucose.(59)

Five prospective studies evaluated insulin-related IPs and pancreatic cancer risk. Three studies from Europe and the USA (736 cases) reported estimates higher than 1.(49,56,60) Among three studies on circulating C-peptide, two reported estimates higher than 1 but with confidence intervals including the null, while one study observed a significant inverse association. (55,57,61)

Two MR studies consistently supported a positive association for genetically-proxied fasting insulin and pancreatic cancer risk. A two-sample MR analysis in European populations (7,110 cases, 7,264 controls), reported an OR of 1.66 (95% CI: 1.03–3.93) per 1-SD increase in fasting insulin.(59) Similarly, a one-sample MR study in the China Kadoorie Biobank found an OR of 1.90 (95% CI: 1.28–2.83).(62)

Out of 514 studies identified on IPs and endometrial cancer incidence, 11 met the inclusion criteria (Supplementary Table 4; Figure 3 and 4), with one additional study identified through reference screening.

Six prospective studies from Europe and the USA (19,157 cases) out of seven reported risk estimates above one when comparing the highest vs. the lowest glucose categories (RR varying between 1.25 and 2.00 with CIs excluding the null for 5 studies).(63–69) A study from South Korea (6,797 cases) also found a positive association between elevated FBG (≥100 mg/dL) and endometrial cancer risk (HR: 1.18, 95% CI: 1.12–1.25).(70)

In contrast, two MR analyses found no strong evidence of an association between genetically-proxied blood glucose levels and endometrial cancer risk (OR per 1 mmol/L increment: 0.95, 95% CI: 0.79–1.14; OR per 1-SD: 1.00, 95% CI: 0.67–1.50).(71,72)

Four prospective cohort studies from Europe and the USA (849 cases) assessed insulin-related IPs and consistently reported risk estimates above 1 for the highest vs. lowest quartile of either fasting insulin, C-peptide or HOMA-IR levels with RR varying between 1.29 and 4.40 but 4 Cis out of 6 including the null.(65,67,69,73)

Two MR analyses supported a positive association between genetically-proxied fasting insulin levels and endometrial cancer risk, reporting ORs of 2.34 (95% CI: 1.06–5.14) per genetically predicted SD increase and 3.93 (95% CI: 2.29–6.74) per 1 natural log–transformed pmol/L increase. (71,72)

Regarding studies on IPs and colorectal cancer incidence, out of 2,600 abstracts, 53 studies met the inclusion criteria (Supplementary Table 5; Figures 5-8).

A total of 30 publications investigated the association between glucose levels and colorectal (CRC), colon, or rectal cancer risk (Supplementary Table 5; Figures 5 and 6). Overall, most studies reported positive associations between glucose levels and CRC risk, with effect estimates ranging from 1.03 to 2.20 when comparing the highest versus lowest categories although the CIs included the null for approximately half of these estimates.(40,74–95) Only three studies reported estimates below 1, with CIs crossing the null. (85,95,96) Similarly, 9 studies out of 10 reported effect estimates ranging from 1.14 to 2.18 between HbA1c highest vs. lowest levels (52,76,92,97–102) while one study reported an RR of 0.83 (95% CI: 0.52-1.33).(103)

For colon cancer, seven studies reported positive associations with glucose levels (74,85–88,104,105) and two reported an inverse association (50,85). Evidence for HbA1c was mixed; Five studies reported positive associations between HbA1c and colon cancer (52,97,99,101,102) and one an estimate below 1 but with a confidence interval including the null (103).

For rectal cancer, eight studies reported a positive association with glucose (77,87,104–106) while one reported an inverse association in men only (50,74,88,106,107). Evidence for HbA1c and rectal cancer was inconsistent, with three estimates above 1 (52,97,99) and two below 1 (102,103).

MR studies provided largely null evidence for glucose. Murphy et al. reported no evidence of association between CRC and genetically-proxied fasting glucose (OR: 1.04; 95% CI: 0.88–1.23) or 2-hour glucose (OR: 1.02; 95% CI: 0.86–1.21), but observed a modest positive association for HbA1c (OR: 1.09; 95% CI: 1.00–1.19), which was stronger for rectal cancer (OR: 1.19; 95% CI: 1.06–1.33). (108) Hanyuda et al. similarly found no association for glucose or HbA1c in East Asian populations.(109)

Ten prospective studies investigated circulating insulin and CRC risk (71 to 1,010 cases) (Supplementary Table 5; Figure 7 and 8). Eight studies reported positive associations, with confidence intervals that included the null for six of them (74,79,86,89,90,92,96,110), and two studies reported estimates below 1.(102,111) No consistent associations were observed for colon cancer, with estimates ranging between 0.80 and 1.28.(74,86,96,102,110) For rectal cancer, two studies showed inconsistent results.(74,110)

Studies on HOMA-IR showed a similar pattern compared to insulin. (74,86,89,90,92,96)

Out of height prospective studies that evaluated circulating C-peptide and CRC six reported positive associations, with risk estimates ranging from 1.17 to 3.20 (112–117), while four (945 cases) reported estimates below 1 (76,98,113,115). For colon cancer, five studies (113–117) showed positive associations with risk ranging from 1.48 to 3.96, while 1 study reported an estimate below 1. For rectal cancer risk, no consistent associations were observed, with estimates ranging from 0.46 to 2.20.(115–117)

MR analyses supported a role for insulin: Murphy et al. reported a positive association between genetically predicted fasting insulin and CRC risk (OR: 1.65; 95% CI: 1.15–2.36). (108) Hanyuda et al. found a non-significant positive association between C-peptide and CRC risk (OR: 1.47; 95% CI: 0.97–2.24). (109)

A total of 826 articles were screened in relation to IPs and ovarian cancer incidence, of which six met the inclusion criteria (Supplementary Table 6; Figure 9).

Three studies assessing glucose levels and ovarian cancer risk (1,333 cases) reported heterogeneous findings, with risk estimates ranging from 0.59 to 1.27.(63,65,118) One study on HbA1c (1,059 cases) reported an inverse association (RR 0.74, 95% CI: 0.62–0.89).(119)

For insulin-related phenotypes, one study in postmenopausal women (120 cases) found no significant association between fasting insulin or HOMA-IR levels and ovarian cancer risk.(65) Another study (132 cases) similarly reported no association for C-peptide (OR 0.89, 95% CI: 0.44–1.83).(73)

One MR study combining data from up to 5,567 diabetes-free individuals and 199,741 UK Biobank participants found no association between fasting insulin and ovarian cancer risk (IVW OR: 0.997; 95% CI: 0.987–1.006).(120)

Out of 715 articles screened in relation to IPs and kidney cancer incidence, 6 met the inclusion criteria (Supplementary Table 7, Figure 10).

Among three prospective studies comparing glucose levels and kidney cancer risk, two reported an HR higher than 1; however, all confidence intervals included the null.(121–123) A UK Biobank study similarly found no association for HbA1c, reporting a hazard ratio of 1.09 (95% CI: 0.92–1.31) for ≥42 vs. <42 mmol/mL.(124)

For insulin-related markers, a nested-case control study reported no association between C-peptide levels and kidney cancer risk (OR 1.08, 95% CI: 0.89–1.30 per 1-SD increment).(125)

In a two-sample MR analysis (10,784 cases, 20,406 controls), no association was observed between fasting glucose and kidney cancer risk (OR 0.92, 95% CI: 0.76–1.12 per 1-SD).(126) In contrast, higher fasting insulin was associated with increased kidney cancer risk ( OR: 1.82; 95% CI: 1.30–2.55 per 1-SD).(126)

Regarding IPs and breast cancer incidence, our analysis primarily built on individual prospective cohorts identified in the systematic review conducted by Drummond et al., who investigated glucose- and insulin-related phenotypes in relation to breast cancer (BC) risk.(127) As their search ended in March 2021, we updated it to identify more recent studies. Out of 689 screened articles, three additional prospective cohort studies and three MR studies were included (Supplementary Table 8).

Three cohorts reported no strong evidence of an association between FBG and postmenopausal BC: WHI- HR: 1.14 per mg/dL, 95%CI=0.60-2.16; ORDET- RR: 1.6 per ng/ml, 95%CI=0.59-4.46, IBIS-II-OR: 1.37 95% CI: 0.74–2.52 (128); however, ORDET observed a positive association with premenopausal BC (RR: 2.76 per mg/dL, 95%CI=1.18-6.46).(128–130)

For HbA1c, one study reported no significant association in premenopausal women, and two studies in postmenopausal women suggested a modest inverse association.(127)

Among three MR studies identified by Drummond et al., two found no effect of genetically predicted fasting glucose on BC risk (OR = 1.03 per mmol/L, 95% CI = 0.85–1.25; OR = 1.06 per mmol/L, 95% CI = 0.95–1.17)(107,131), with one reporting an inverse association in postmenopausal women (HR = 0.59, 95% CI = 0.35–0.99)(132). Additional MR studies also found no overall association for glucose and glycated haemoglobin, including by ER status.(133–135)

For fasting insulin, Drummond et al. reported a pooled RR of 1.12 (95%CI:0.30-1.94, I^2^=67%, 3 studies), and three additional cohort studies reported no statistically significant associations (RR ranging from 0.88 to 1.46).(128,136,137)

Four cohort studies on C-peptide yielded a pooled RR of 1.16 (95% CI: 0.93–1.37; I² = 0%), regardless of menopausal status, and an additional study among postmenopausal Australian women (347 cases) reported an RR of 0.91 (95% CI: 0.70–1.18).(137)

For HOMA-IR, one cohort study reported a positive association in postmenopausal women (HR: 2.99, 95% CI: 1.56–5.73), while other studies reported null findings.(128,136)

Two MR studies included in Drummond et al. reported that higher insulin levels were associated with increased BC risk (OR = 1.71 per 1-SD, 95% CI: 1.26–2.31; HR = 1.80 per μIU/ml, 95% CI: 0.18–18.06). Later MR analyses reported weaker positive associations (OR = 1.29, 95% CI: 0.86–1.94(133) and OR = 1.27, 95% CI: 1.03–1.56 (134))One MR on HOMA-IR and postmenopausal BC showed no association (HR = 0.94, 95% CI: 0.81 to 1.08).(127)

Grading of the evidence by the MEC (stage 3) is presented in Table 1. Glucose and insulin-related IPs were evaluated jointly, except in the case of endometrial cancer, as these association results differed significantly.

**Table 1:** Assessment of the strength of evidence for the associations between sugar sweetened beverages (SSB) or artificially sweetened beverages (ASB), IPs, and the indicated cancer sites.

|  |  |  | Exposure to Intermediate phenotypes |  |
| --- | --- | --- | --- | --- |
|  |  |  | SSB | ASB |
|  |  |  | Limited | Limited to Probable |
| Intermediate phenotype to Outcomes | Colon | Limited | Weak | Weak |
|  | Rectal | Limited | Weak | Weak |
|  | Colorectal | Probable | Weak | Weak |
|  | Ovary | Limited | Weak | Weak |
|  | Endometrial | Probable (glucose)<br>Convincing (insulin) | Weak | Weak |
|  | Pancreatic | Probable to Convincing | Weak | Weak |
|  | Kidney | Limited | Weak | Weak |
|  | Breast (overall, pre/post-menopausal) | Limited | Weak | Weak |

Abbreviations: ASB: artificially-sweetened beverages; SSB: sugar-sweetened beverages

The evidence of an association between SSB consumption and glucose/insulin-related IPs was judged as *limited*, whereas the association with ASB was evaluated as *limited* to *probable*.

Regarding the strength of evidence linking IPs with individual cancer sites, most associations were rated as *limited*, except for CRC, pancreatic cancer and endometrial cancer, for which the evidence was judged as *probable* to *convincing*.

Overall, there is weak evidence that insulin/glucose-related IPs operate as a plausible mechanism between ASB or SSB and risk of colorectal, pancreatic, kidney, ovarian, endometrial and breast cancers.

## Discussion

Based on the evidence from human studies of biological processes linking soft drink consumption to cancer risk, the MEC concluded that there was probable to convincing evidence supporting a role of insulin/glucose in cancers of the colorectum, endometrium and pancreas while the evidence was weak for cancers of the ovary, kidney, breast and for colon and rectum cancers separately. However, the available evidence was weak regarding the evidence on SSBs and ASBs consumption and glucose/insulin. Consequently, the MEC concluded that there was *weak* evidence supporting glucose/insulin as a plausible mechanism linking SSB or ASBs consumption to colon, rectal, colorectal, ovarian, kidney, pancreatic, endometrial and breast (overall and by menopausal status) cancers.

This is the first narrative review using the CUP Global methodology to examine mechanistic evidence (insulin and glucose-related phenotypes) linking soft drink consumption to multiple cancer sites.

A key strength of this review is the systematic approach to studying identification, selection, and appraisal, reducing the risk of bias and enhancing reproducibility and transparency. Oversight and guidance from the MEC throughout the review process further strengthened methodological rigor and scientific validity.

However, several limitations should be acknowledged. First, substantial heterogeneity existed across studies of exposure, dietary assessment methods, comparison groups, and timing of measurements, limiting comparability. Some studies were also underpowered, particularly for rarer cancers. Second, soft drink intake in cohort studies was mainly self-reported, introducing potential measurement error and misclassification. Many studies relied on a single time point dietary assessment, which may not reflect long-term consumption. Third, soft drink consumption often co-occurs with other lifestyle factors (e.g. total energy intake, diet quality, smoking, alcohol consumption, physical activity, socioeconomic status, diabetes, and adiposity), and residual confounding, especially by BMI, may persist. Finally, variability in study design and adjustment strategies precluded firm conclusions on dose–response relationships and limited subtype-specific analyses.

From a biological perspective, several plausible mechanisms may link soft drink consumption, hyperglycaemia, and hyperinsulinemia, to cancer (138,139). Elevated insulin can increase IGF-I levels, leading to greater activation of insulin and IGF I receptors, activating downstream PI3K–AKT–mTOR and Ras–MAPK pathways that promote cell proliferation, inhibit apoptosis, and stimulate angiogenesis. In parallel, higher circulating glucose availability and the insulin-driven upregulation of glucose transport may enhance tumor metabolism, oxidative stress, and DNA damage, supporting carcinogenesis. Although these pathways are biologically plausible, the evidence linking soft drink consumption itself to insulin and glucose remains inconsistent. In contrast, associations between intermediate metabolic phenotypes (glucose and insulin) and cancer risk appear more consistent.

Further high-quality, mechanistically oriented research is needed. Future studies should use standardised definitions of soft drink subtypes, improved and repeated dietary assessment, and cancer heterogeneity by subtype. Additional mechanisms, such as visceral adiposity, which may influence carcinogenesis through adipokine dysregulation, should also be explored.(140–143) Longitudinal studies incorporating repeated biomarker measurements and more diverse populations are essential to improve causal inference and generalisability. Broader synthesis approaches, such as umbrella reviews, may also help integrate evidence across intermediate phenotypes.

This work contributed to CUP Global evaluations alongside epidemiological reviews conducted by Imperial College London, informing expert panel conclusions and public health recommendations for cancer prevention. Although human studies currently support a probable positive association between SSB consumption and risk of certain cancers (Jayedi at al, unpublished), the evidence supporting insulin- and glucose-related pathways as underlying mechanisms linking this association is weak. The evidence regarding ASBs remains limited. Future studies should prioritise mechanistic research with longitudinal designs and harmonised, standardised, and repeated assessment of dietary exposures, including soft drinks, as well as of intermediate phenotypes.

## Data Availability

All data produced in the present work are contained in the manuscript

## Acknowledgements

We acknowledge input from the CUP Global Secretariat (some individual members are listed as co-authors): Panagiota Mitrou, Helen Croker, Vanessa Gordon-Dseagu, Sarah Kefyalew, Holly Paden, Elio Riboli, and Christelle Clary for providing overall strategic direction and coordination for the work and for convening and facilitating discussions with the CUP Global Expert Committees and Expert Panel. We acknowledge the foundational work undertaken by the Biological Process Workstream from the CUP transition: Marc Gunter, Stephen Hursting, Sarah Lewis, Kate Guyton, Christian Frezza, Vivien Lund, and Steven Clinton. We acknowledge the contributions of the CUP Global team at Imperial College London.

## IARC disclaimer

Where authors are identified as personnel of the International Agency for Research on Cancer/World Health Organization, the authors alone are responsible for the views expressed in this article and they do not necessarily represent the decisions, policy, or views of the International Agency for Research on Cancer/World Health Organization.

---

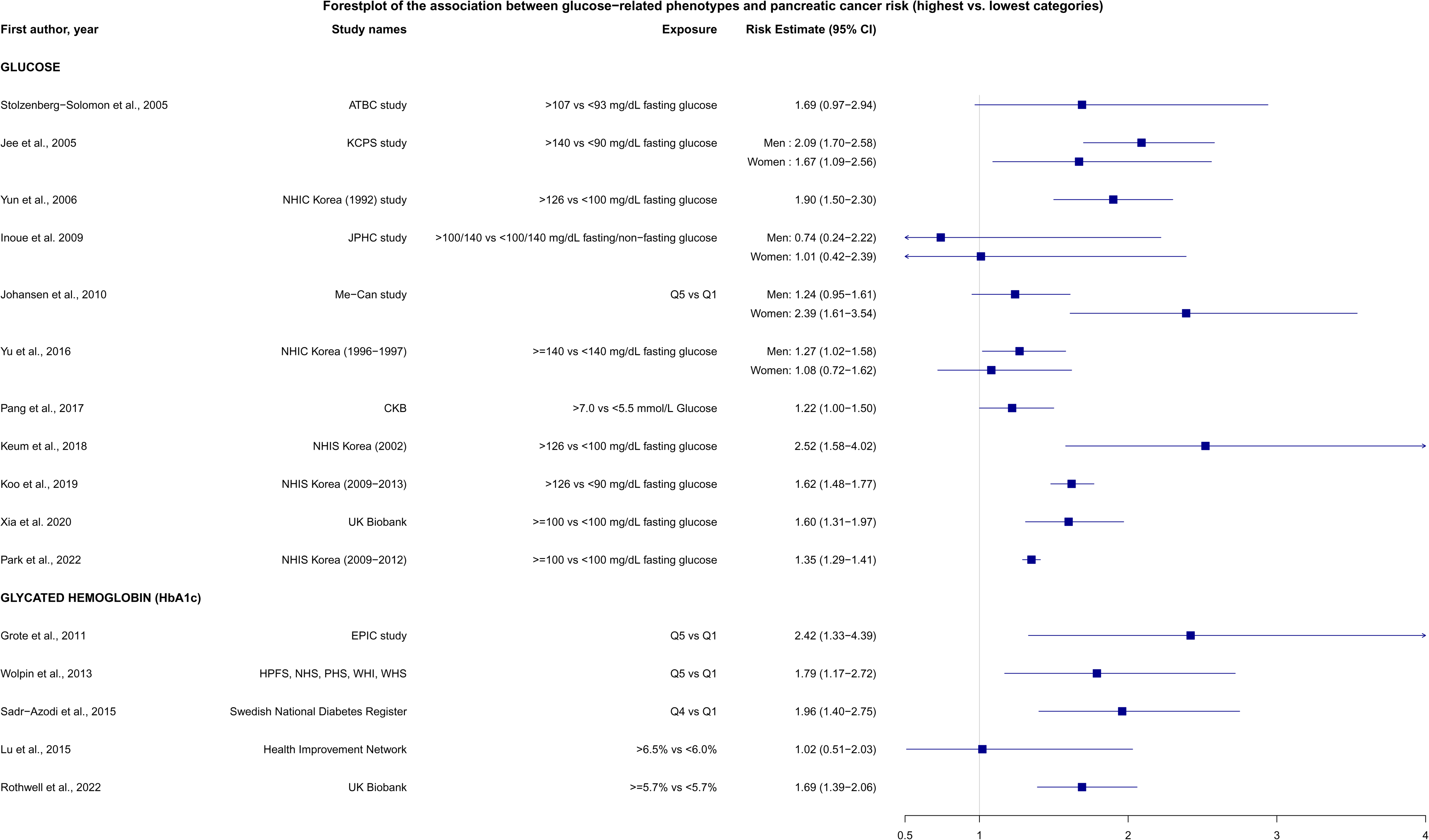

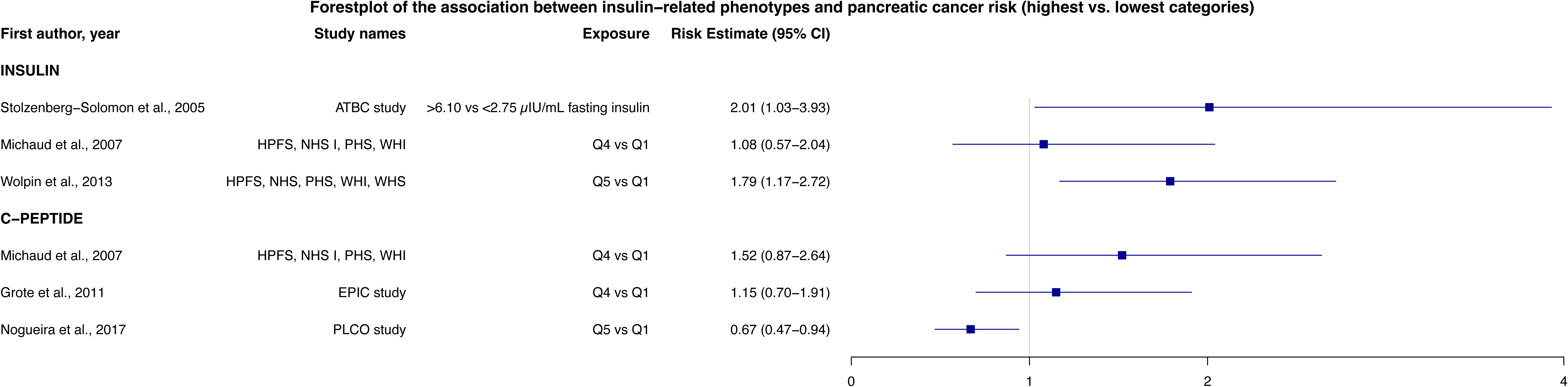

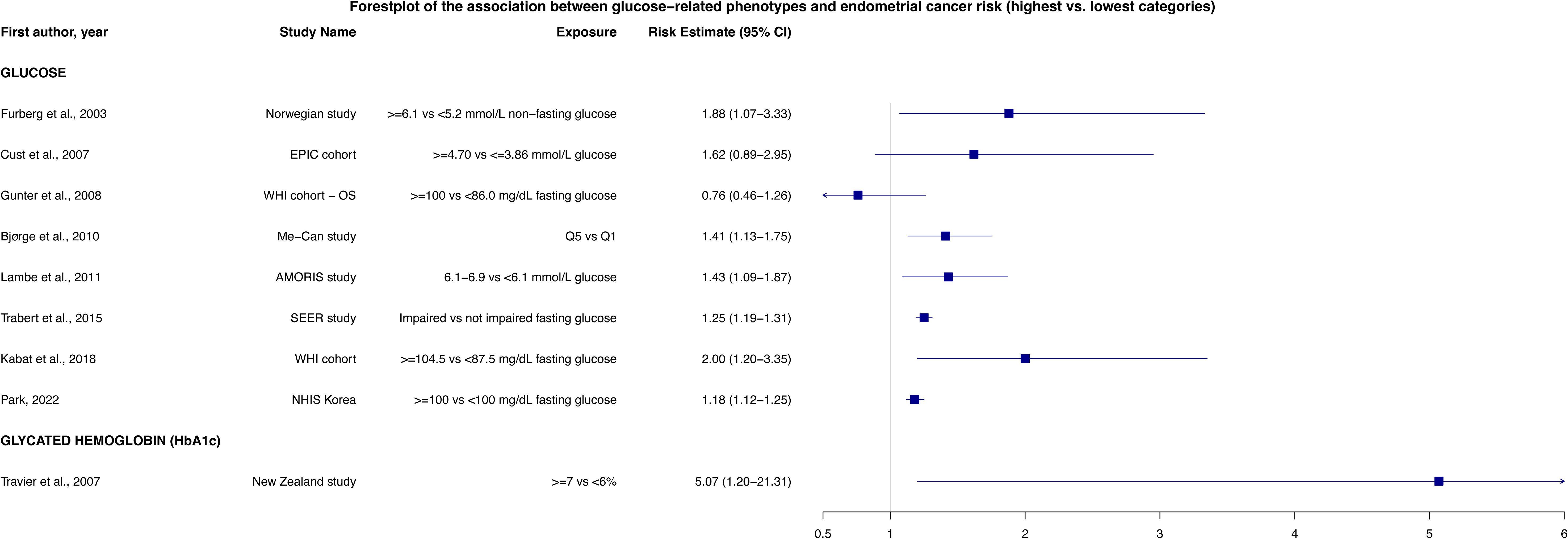

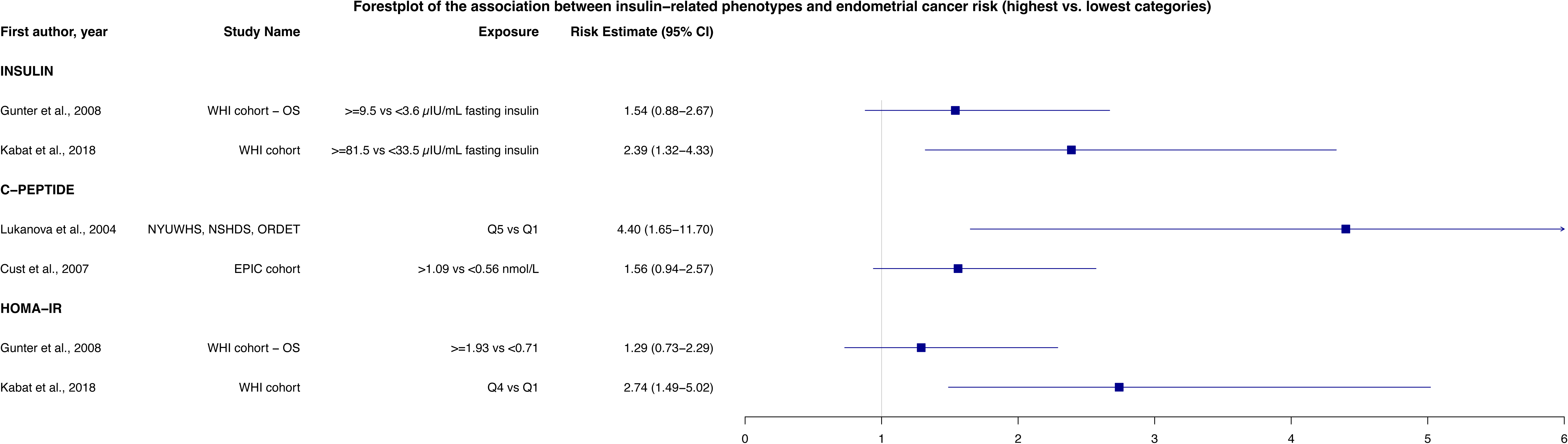

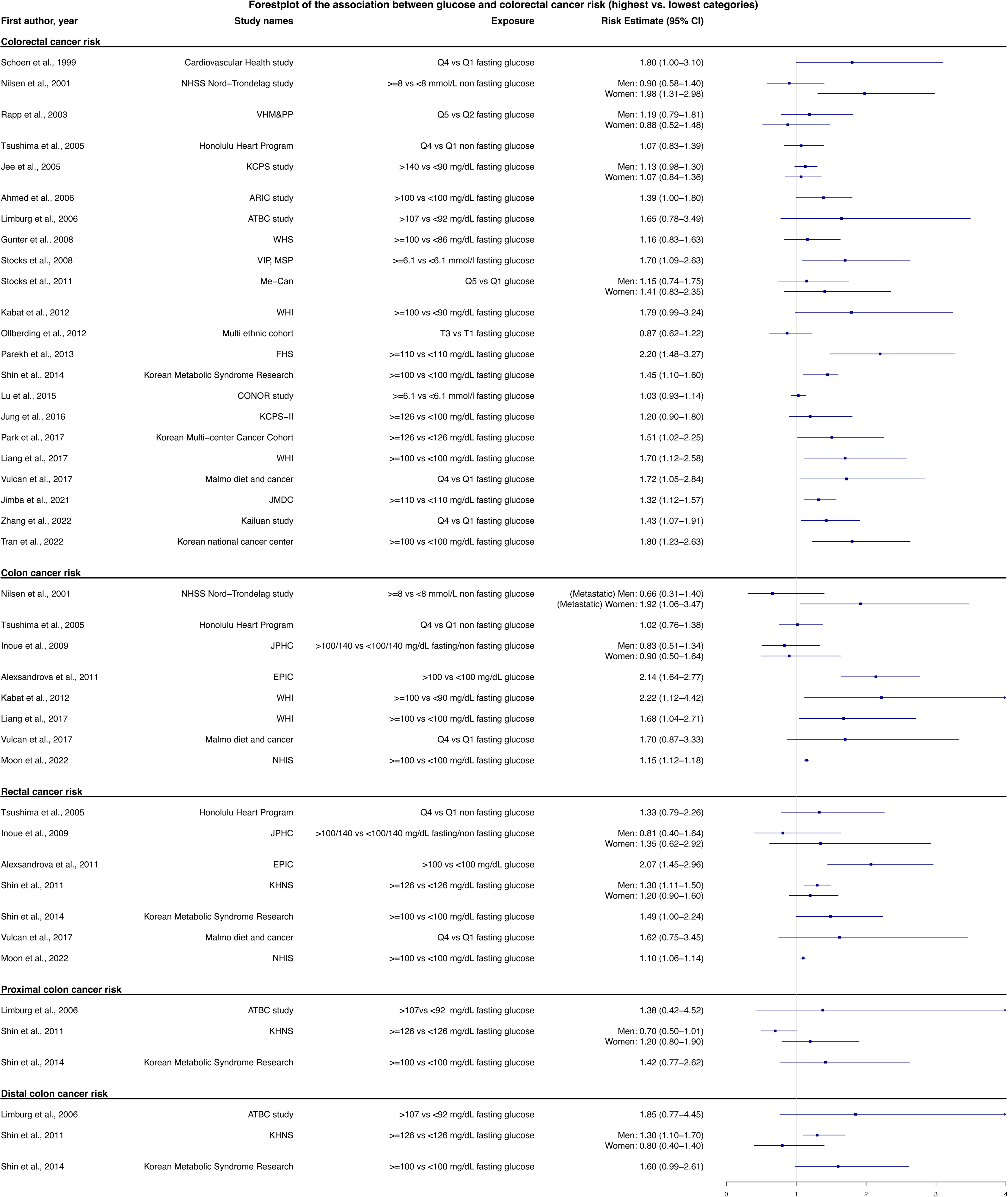

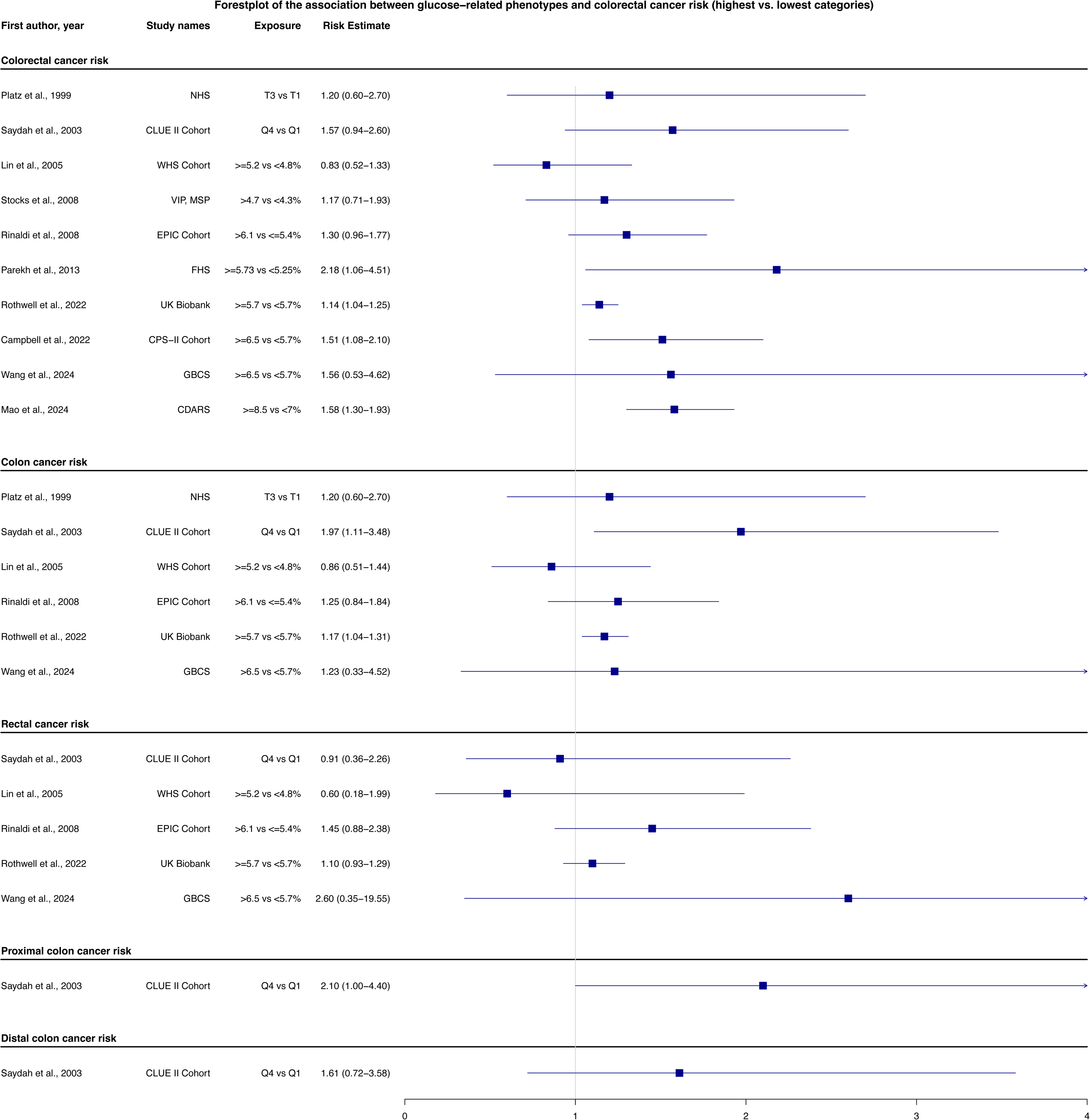

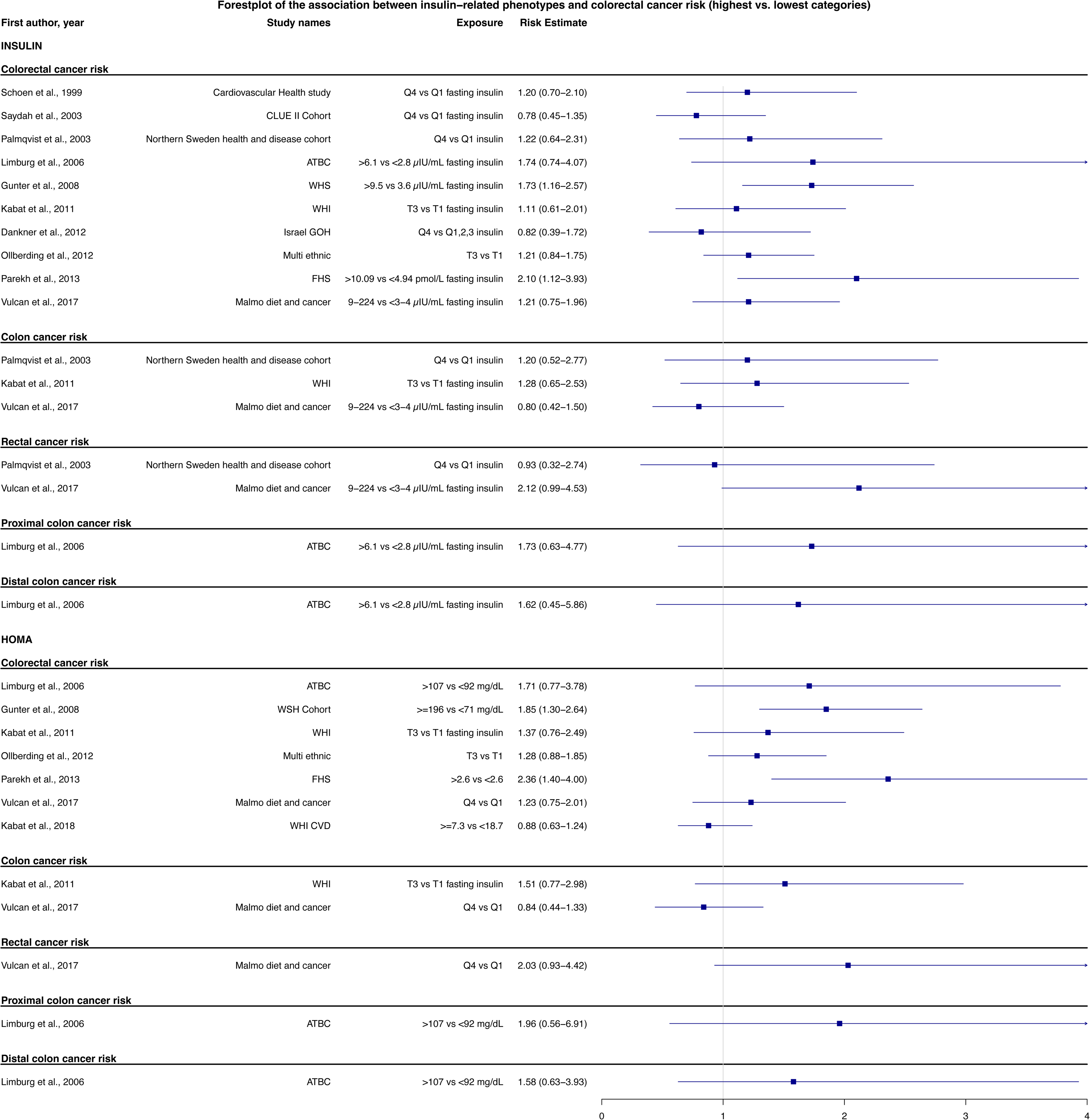

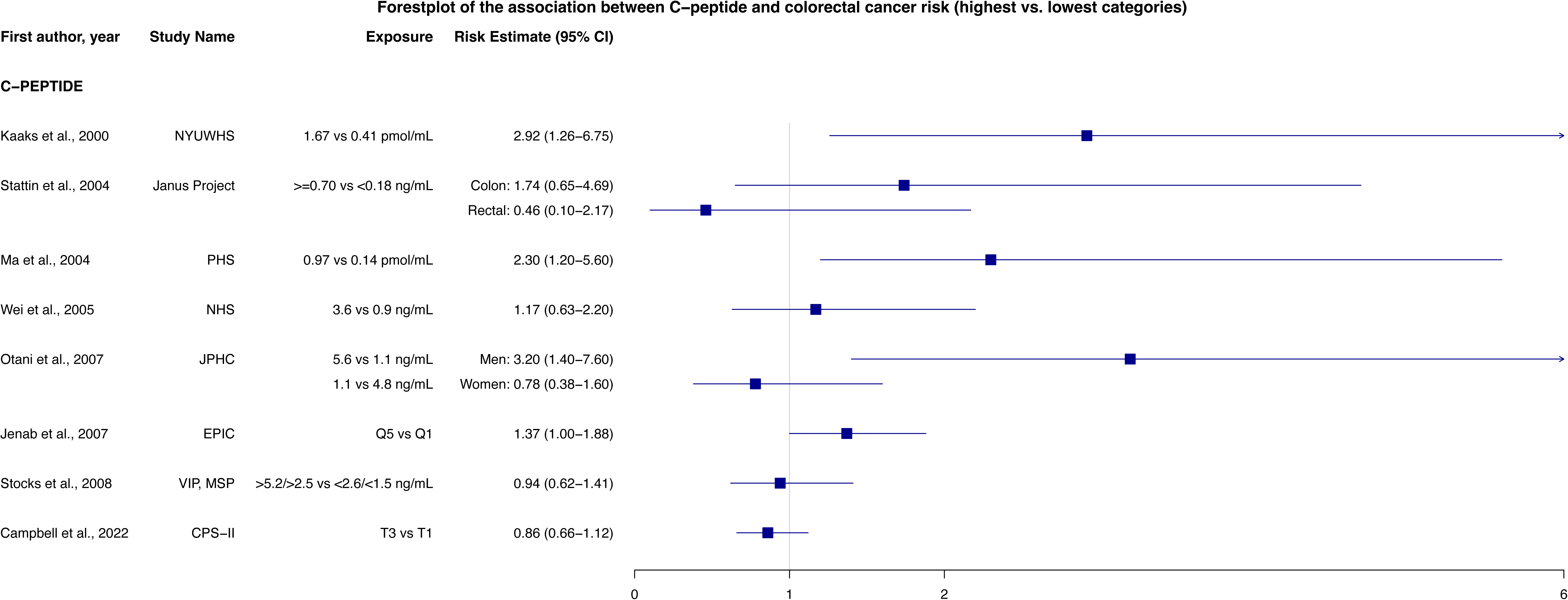

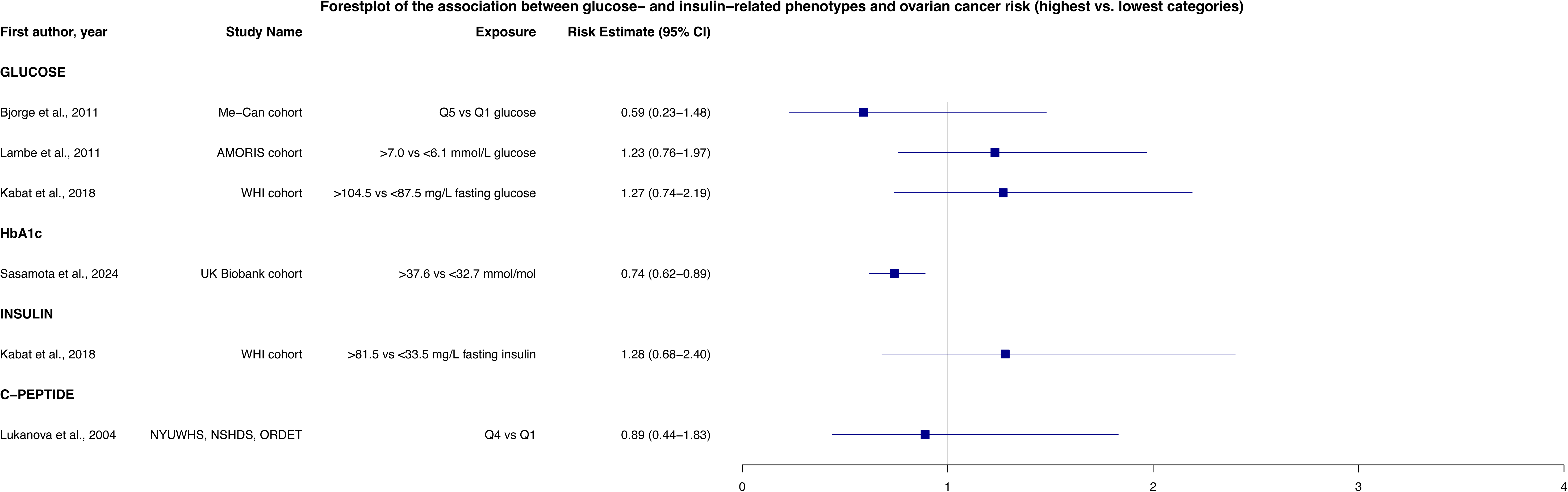

