## supplementary material for "Biological processes linking soft drink consumption with site-specific cancer risk within the Global Cancer Update Programme (CUP Global)"

Fontvieille et al.

### **Supplementary material 1. Definitions**

### *Soft drinks*

Soft drinks refer to non-alcoholic sweetened beverages, which include both SSBs and ASBs. SSBs are drinks sweetened by adding free sugars, such as sucrose, high-fructose corn syrup, or sugars naturally present in honey, syrups, fruit juices, or juice concentrates, and include products such as sodas, sports drinks, energy drinks, sweetened waters, cordials, barley water, and coffee or tea beverages with added sugar. In contrast, ASBs are low- or non-caloric versions of these drinks, sweetened with artificial sweeteners such as aspartame, acesulfame-K, saccharin, sucralose, or neotame, and are often marketed as “diet,” “low sugar,” or “sugar-free” beverages.

### *Glucose- and insulin-related IPs*

The glucose-related biomarkers considered included fasting blood glucose (FBG), 2-hour postprandial (2hpp) glucose, glycoalbumin and glycated haemoglobin (HbA1c), the latter being an indicator of long-term glycaemic control that reflects average blood glucose levels over the previous 2 to 3 months. Insulin-related IPs included insulin, C-peptide (a marker of pancreatic insulin secretion), and the homeostasis model assessment of insulin resistance (HOMA-IR), an index calculated from fasting glucose and fasting insulin concentrations to estimate insulin resistance, HOMA-B and other insulin-related indices.

### **Supplementary Material 2: PubMed search strings and MeSH terms for TEMMPO for Stage 1**

| **PubMed search strings for**  **(E AND IPs) OR (IPs AND O)** | (("Carbonated Beverages"[MeSH Terms] OR "Artificially Sweetened Beverages"[MeSH Terms] OR "Sugar-Sweetened Beverages"[MeSH Terms] OR "Fruit and Vegetable Juices"[MeSH Terms] OR "beverage*"[Text Word] OR "sugar* drink*"[Text Word] OR "sugar containing drink*"[Text Word] OR "carbonated drink*"[Text Word] OR "soft drink*"[Text Word] OR "energy drink*"[Text Word] OR "sweet* drink*"[Text Word] OR "nonalcoholic drink*"[Text Word] OR "non-alcoholic drink*"[Text Word] OR "Soda"[Text Word] OR "juice*"[Text Word] OR "sugar* food*"[Text Word] OR "Added sugar"[Text Word] OR "fruit drink*"[Text Word])  AND  ("inflammation"[MeSH Terms] OR "cytokines"[MeSH Terms] OR "chemokines"[MeSH Terms] OR "immune system"[MeSH Terms] OR "insulin"[MeSH Terms] OR "Insulin-Like Growth Factor I"[MeSH Terms] OR "glucose"[MeSH Terms] OR "c peptide"[MeSH Terms] OR "hyperinsulinism"[MeSH Terms] OR "lipids"[MeSH Terms] OR "Fatty Acids"[MeSH Terms] OR "dyslipidemias"[MeSH Terms] OR "hyperlipidemias"[MeSH Terms] OR "hypercholesterolemia"[MeSH Terms] OR "hypertriglyceridemia"[MeSH Terms] OR "steroids"[MeSH Terms] OR "prolactin"[MeSH Terms] OR "Gonadal Hormones"[MeSH Terms] OR "Thyroid Hormones"[MeSH Terms] OR "melatonin"[MeSH Terms] OR "Endocrine Disruptors"[MeSH Terms] OR "metabolomics"[MeSH Terms] OR "metabolome"[MeSH Terms] OR "Amino acids"[MeSH Terms] OR "microbiota"[MeSH Terms] OR "oxidative stress"[MeSH Terms] OR "antioxidants"[MeSH Terms] OR "reactive oxygen species"[MeSH Terms] OR "epigenesis, genetic"[MeSH Terms] OR "epigenomics"[MeSH Terms] OR "histones"[MeSH Terms] OR "dna methylation"[MeSH Terms] OR "micrornas"[MeSH Terms] OR "dna damage"[MeSH Terms] OR "dna repair"[MeSH Terms] OR "mutagens"[MeSH Terms] OR "oncogenes"[MeSH Terms] OR "genomic instability"[MeSH Terms]))  OR  (("inflammation"[MeSH Terms] OR "cytokines"[MeSH Terms] OR "chemokines"[MeSH Terms] OR "immune system"[MeSH Terms] OR "insulin"[MeSH Terms] OR "Insulin-Like Growth Factor I"[MeSH Terms] OR "glucose"[MeSH Terms] OR "c peptide"[MeSH Terms] OR "hyperinsulinism"[MeSH Terms] OR "lipids"[MeSH Terms] OR "Fatty Acids"[MeSH Terms] OR "dyslipidemias"[MeSH Terms] OR "hyperlipidemias"[MeSH Terms] OR "hypercholesterolemia"[MeSH Terms] OR "hypertriglyceridemia"[MeSH Terms] OR "steroids"[MeSH Terms] OR "prolactin"[MeSH Terms] OR "Gonadal Hormones"[MeSH Terms] OR "Thyroid Hormones"[MeSH Terms] OR "melatonin"[MeSH Terms] OR "Endocrine Disruptors"[MeSH Terms] OR "metabolomics"[MeSH Terms] OR "metabolome"[MeSH Terms] OR "Amino acids"[MeSH Terms] OR "microbiota"[MeSH Terms] OR "oxidative stress"[MeSH Terms] OR "antioxidants"[MeSH Terms] OR "reactive oxygen species"[MeSH Terms] OR "epigenesis, genetic"[MeSH Terms] OR "epigenomics"[MeSH Terms] OR "histones"[MeSH Terms] OR "dna methylation"[MeSH Terms] OR "micrornas"[MeSH Terms] OR "dna damage"[MeSH Terms] OR "dna repair"[MeSH Terms] OR "mutagens"[MeSH Terms] OR "oncogenes"[MeSH Terms] OR "genomic instability"[MeSH Terms])  AND  #("colorectal neoplasms"[MeSH Terms] OR “colon neoplasms"[MeSH Terms]))  #("endometrial neoplasms"[MeSH Terms]))  #("pancreatic neoplasms"[MeSH Terms])) |
| --- | --- |
| **MeSH Terms for TEMMPO:** | Exposure:  Carbonated Beverages ; Artificially Sweetened Beverages ; Sugar-Sweetened Beverages ; Fruit and Vegetable Juices   - *Artificially Sweetened Beverages: could not be found* - *Sugar-Sweetened Beverages: could not be found*   Mediators:  Inflammation ; cytokines ; chemokines ; immune system ; Insulin ; Insulin-Like Growth Factor I ; Glucose ; C-peptide ; hyperinsulinism ; Lipids; Fatty Acids ; Dyslipidemias; Hyperlipidemias ; Hypercholesterolemia ; Hypertriglyceridemia ; Steroids ; Prolactin; Gonadal Hormones ; Thyroid Hormones ; Melatonin ; Endocrine Disruptors ; Metabolomics ; Metabolome ; Amino acids ; Microbiota ; Oxidative stress ; Antioxidants ; reactive oxygen species ; epigenesis, genetic ; epigenomics ; histones ; DNA Methylation ; MicroRNAs ; DNA damage ; DNA repair ; Mutagens ; Oncogenes ; genomic instability  Outcomes:  colorectal neoplasms; colonic neoplasms  endometrial neoplasms  pancreatic neoplasms |

Abbreviations: E: Exposure; IPs: Intermediate phentoypes; MeSH: Medical subject headings; O: Outcomes

### **Supplementary Material 3: PubMed search strings for stage 2**

|  | **PubMed search strings** |
| --- | --- |
| **E & IPs** | (("Carbonated Beverages"[MeSH Terms] OR "Artificially Sweetened Beverages"[MeSH Terms] OR "Sugar-Sweetened Beverages"[MeSH Terms] OR "Fruit and Vegetable Juices"[MeSH Terms] OR "beverage*"[Text Word] OR "sugar* drink*"[Text Word] OR "sugar containing drink*"[Text Word] OR "carbonated drink*"[Text Word] OR "soft drink*"[Text Word] OR "energy drink*"[Text Word] OR "sweet* drink*"[Text Word] OR "nonalcoholic drink*"[Text Word] OR "non-alcoholic drink*"[Text Word] OR "Soda"[Text Word] OR "juice*"[Text Word] OR "sugar* food*"[Text Word] OR "Added sugar"[Text Word] OR "fruit drink*"[Text Word])  AND  ("Insulin"[MeSH Terms] OR "C-Peptide"[MeSH Terms] OR "Hyperinsulinism"[MeSH Terms] OR "Glucose"[MeSH Terms] OR "Insulin Resistance"[MeSH Terms] OR "Glycated Hemoglobin"[MeSH Terms] OR "Blood Glucose"[MeSH Terms] OR "Hyperglycemia"[MeSH Terms] OR "glucose metabolism"[Text Word] OR "fasting glucose"[Text Word] OR "glycemic response"[Text Word] OR "insulin"[Text Word] OR "HOMA-IR"[Text Word] OR "HbA1c"[Text Word])  AND  ("Humans"[MeSH Terms]) |
| **IPs & PC** | ("Insulin"[MeSH Terms] OR "C-Peptide"[MeSH Terms] OR "Hyperinsulinism"[MeSH Terms] OR "Glucose"[MeSH Terms] OR "Insulin Resistance"[MeSH Terms] OR "Glycated Hemoglobin"[MeSH Terms] OR "Blood Glucose"[MeSH Terms] OR "Hyperglycemia"[MeSH Terms] OR "glucose metabolism"[Text Word] OR "fasting glucose"[Text Word] OR "glycemic response"[Text Word] OR "insulin"[Text Word] OR "HOMA-IR"[Text Word] OR "HbA1c"[Text Word])  AND  ("pancreatic neoplasms"[MeSH Terms])) |
| **IPs & EC** | ("Insulin"[MeSH Terms] OR "C-Peptide"[MeSH Terms] OR "Hyperinsulinism"[MeSH Terms] OR "Glucose"[MeSH Terms] OR "Insulin Resistance"[MeSH Terms] OR "Glycated Hemoglobin"[MeSH Terms] OR "Blood Glucose"[MeSH Terms] OR "Hyperglycemia"[MeSH Terms] OR "glucose metabolism"[Text Word] OR "fasting glucose"[Text Word] OR "glycemic response"[Text Word] OR "insulin"[Text Word] OR "HOMA-IR"[Text Word] OR "HbA1c"[Text Word])  AND  ("endometrial neoplasms"[MeSH Terms])) |
| **IPs & CRC** | ("Insulin"[MeSH Terms] OR "C-Peptide"[MeSH Terms] OR "Hyperinsulinism"[MeSH Terms] OR "Glucose"[MeSH Terms] OR "Insulin Resistance"[MeSH Terms] OR "Glycated Hemoglobin"[MeSH Terms] OR "Blood Glucose"[MeSH Terms] OR "Hyperglycemia"[MeSH Terms] OR "glucose metabolism"[Text Word] OR "fasting glucose"[Text Word] OR "glycemic response"[Text Word] OR "insulin"[Text Word] OR "HOMA-IR"[Text Word] OR "HbA1c"[Text Word])  AND  ("colorectal neoplasms"[MeSH Terms] OR “colon neoplasms"[MeSH Terms])) |
| **IPs & OC** | ("Insulin"[MeSH Terms] OR "C-Peptide"[MeSH Terms] OR "Hyperinsulinism"[MeSH Terms] OR "Glucose"[MeSH Terms] OR "Insulin Resistance"[MeSH Terms] OR "Glycated Hemoglobin"[MeSH Terms] OR "Blood Glucose"[MeSH Terms] OR "Hyperglycemia"[MeSH Terms] OR "glucose metabolism"[Text Word] OR "fasting glucose"[Text Word] OR "glycemic response"[Text Word] OR "insulin"[Text Word] OR "HOMA-IR"[Text Word] OR "HbA1c"[Text Word])  AND  ("ovarian neoplasms"[MeSH Terms])) |
| **IPs & KC** | ("Insulin"[MeSH Terms] OR "C-Peptide"[MeSH Terms] OR "Hyperinsulinism"[MeSH Terms] OR "Glucose"[MeSH Terms] OR "Insulin Resistance"[MeSH Terms] OR "Glycated Hemoglobin"[MeSH Terms] OR "Blood Glucose"[MeSH Terms] OR "Hyperglycemia"[MeSH Terms] OR "glucose metabolism"[Text Word] OR "fasting glucose"[Text Word] OR "glycemic response"[Text Word] OR "insulin"[Text Word] OR "HOMA-IR"[Text Word] OR "HbA1c"[Text Word])  AND  ("kidney neoplasms"[MeSH Terms])) |
| **IPs & BC** | ("Insulin"[MeSH Terms] OR "C-Peptide"[MeSH Terms] OR "Hyperinsulinism"[MeSH Terms] OR "Glucose"[MeSH Terms] OR "Insulin Resistance"[MeSH Terms] OR "Glycated Hemoglobin"[MeSH Terms] OR "Blood Glucose"[MeSH Terms] OR "Hyperglycemia"[MeSH Terms] OR "glucose metabolism"[Text Word] OR "fasting glucose"[Text Word] OR "glycemic response"[Text Word] OR "insulin"[Text Word] OR "HOMA-IR"[Text Word] OR "HbA1c"[Text Word])  AND  ("breast neoplasms"[MeSH Terms])) |

Abbreviations: BC: Breast Cancer; CRC: Colorectal cancer; E: Exposure; EC: Endometrial cancer; IPs: Intermediate phentoypes; KC: Kidney cancer; MeSH: Medical subject headings;OC: Ovarian cancer; PC: Pancreatic cancer

**Supplementary Material 4: Inclusion and exclusion criteria**

| **E & IPs** | Inclusion criteria:  Population: Studies involving adults  Exposure: Soft drinks; sugar sweetened beverages and artificially sweetened beverages combined sugar sweetened beverages; artificially sweetened beverages; fruit and vegetable juices; fruit juices; vegetable juices; specific types of soft drinks, such as carbonated or non-carbonated, sugar sweetened or artificially sweetened, soda or fruit drinks, pure fruit juices, specific fruit juices (orange juice, citrus juice, apple juice, grapefruit juice), or tomato juice  Comparison: No consumption of soft drinks, or consumption of water/other beverages  Outcomes: Changes in glucose-related phenotypes: glucose, 2hpp glucose, glycated haemoglobin and insulin-related phenotypes: insulin, C-peptide, insulin resistance indices (HOMA-insulin sensitivity, Matsuda Insulin Sensitivity Index and HOMA-IR) and insulin secretion indices (HOMA-B, insulinogenic index, disposition index, Stumvoll first- and second-phase indices, and the ratio of insulinemia AUC to glycemia AUC)  Study design: Randomised controlled trials (RCTs), prospective cohort studies  Sample size: ≥50 (for RCTs)  Exposure duration: Interventions administered over an extended period, rather than a single exposure in one day (for RCTs)  Language: Studies published in English  Exclusion criteria:  Population: Children or adolescents  Study design: Cross-over trials, randomised cross-over trials, intervention studies, retrospective studies, such as case-control and cross-sectional designs, reports, narrative reviews and meta-analysis |
| --- | --- |
| **IPs & O** | Inclusion criteria:  Population: Studies involving adults  Exposure: glucose- and insulin-related phenotypes (Glucose, glycoalbumin, glycated haemoglobin, insulin, HOMA-IR, HOMA, C-peptide)  Comparison: varying levels of these biomarkers  Outcomes: Endometrial, colorectal, pancreatic incidence  Study design: Prospective cohort studies, including nested case-control and case-cohort studies as well as Mendelian Randomization studies  Language: Studies published in English  Exclusion criteria:  Population: Children or adolescents  Study design: Retrospective studies, such as case-control and cross-sectional designs, reports, narrative reviews and meta-analysis |

Abbreviations: E: Exposure; IPs: Intermediate phentoypes; O: Outcomes

**Supplementary Material 5: list of data extracted**

- PubMed ID
- first author
- year of publication
- country
- study design
- sample size
- duration of follow-up
- participant age
- relevant population-specific characteristics
- IPs considered
- specific biomarkers assessed
- method of exposure measurement
- nature of the exposure comparison (e.g., continuous or categorical),
- adjustment factors
- main results

For RCTs, comparison group and duration of exposure were also extracted.

For studies on cancer outcomes, we additionally extracted the number of cancer cases.

**Supplementary Material 6: Assessing the plausibility of the overall mechanism**

| **E&IP**  **IP&0** | **Convincing** | **Probable** | **None or limited** |
| --- | --- | --- | --- |
| **Convincing** | **Strong** | **Strong** | **Weak or no evidence available** |
| **Probable** | **Strong** | **Moderate** | **Weak or no evidence available** |
| **None or limited** | **Weak or no evidence available** | **Weak or no evidence available** | **Weak or no evidence available** |

Abbreviations: E: exposure; IPs: Intermediate phenotypes; O: outcomes

**Guiding criteria:**

**Convincing evidence:** evidence from more than one study type, including RCTs, Mendelian randomization studies, mediation analysis studies, cohort studies (at least 2 independent cohort studies), or pooled or meta-analysis of these studies; good quality studies; consistency across the studies and study types; large magnitude of effect or biological gradient (dose-response). Note that good-quality studies exclude with confidence the possibility that the observed association results from random or systematic error, including confounding, measurement error and selection bias. This decision is left to the judgement of the MEC based on the study-level information provided, including study design, sample size, exposure assessment, outcome ascertainment and adjustment factors.

**Probable evidence:** evidence that is stronger than “limited” but weaker than “convincing”

**None or limited evidence:** too few or no studies available; inconsistency of direction of effect; poor quality of studies (for example, lack of adjustment for important confounders)

**Supplementary Figure 1: Bubble plots of the automated searches for soft drinks consumption and pancreatic, colorectal or endometrial cancer incidence**

| **Pancreatic cancer** | 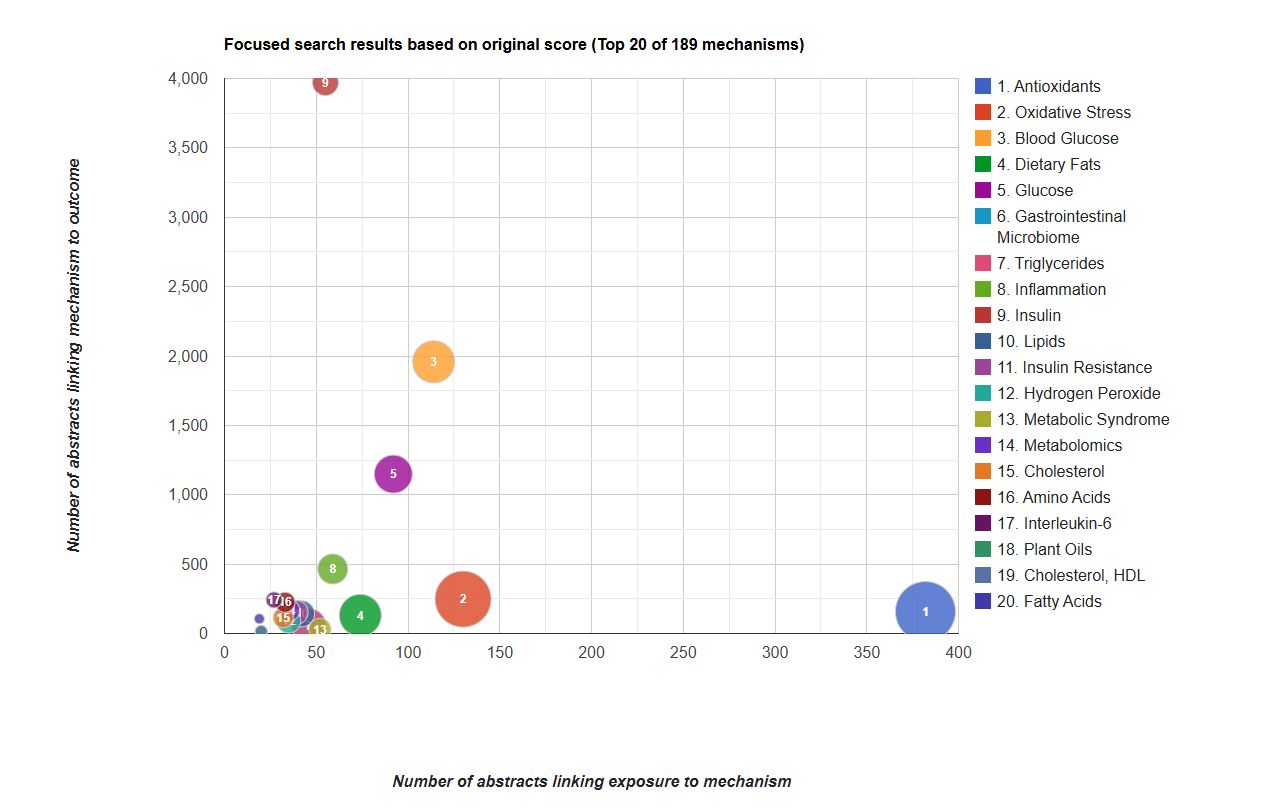 |
| --- | --- |
| **Colorectal cancer** | 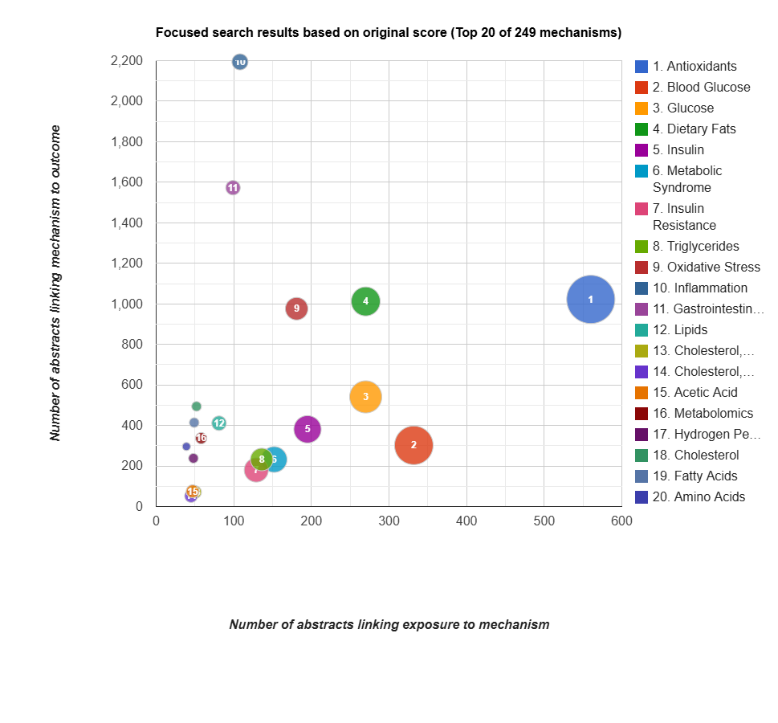 |
| **Endometrial cancer** | 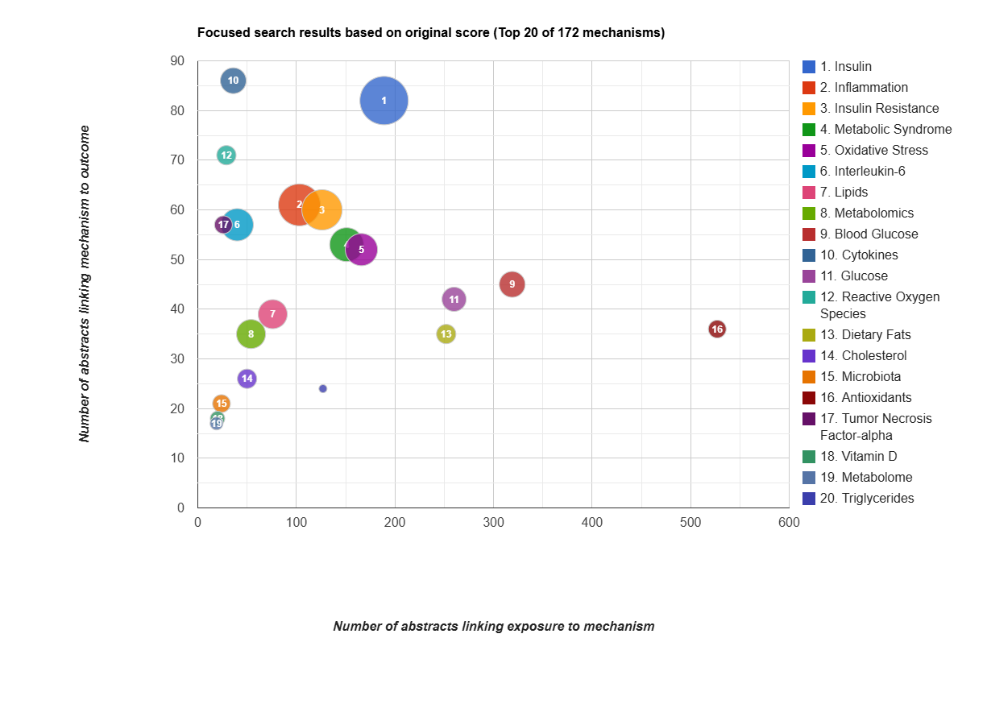 |
| Notes: Based on the MeSH used as described in Annex 1, *Artificially Sweetened Beverages and Sugar-Sweetened Beverages w*ere not found in the TEMMPO websites. | |

**Supplementary table 1. Main characteristics and results of prospective cohorts evaluating associations between soft drinks and insulin- or glucose-related intermediate phenotypes**

**Supplementary table 2. Main characteristics and results of randomised controlled trials (RCTs) evaluating associations between soft drinks and insulin- or glucose-related intermediate phenotypes**

**Supplementary table 3. Main characteristics and results of prospective cohorts evaluating associations between insulin- or glucose-related intermediate phenotypes and risk of pancreatic cancer**

**Supplementary table 4. Main characteristics and results of prospective cohorts evaluating associations between insulin- or glucose-related intermediate phenotypes and risk of colorectal cancer**

**Supplementary table 5. Main characteristics and results of prospective cohorts evaluating associations between insulin- or glucose-related intermediate phenotypes and risk of endometrial cancer**

**Supplementary table 6. Main characteristics and results of prospective cohorts evaluating associations between insulin- or glucose-related intermediate phenotypes and risk of ovarian cancer**

**Supplementary table 7. Main characteristics and results of prospective cohorts evaluating associations between insulin- or glucose-related intermediate phenotypes and risk of kidney cancer**

**Supplementary table 8. Main characteristics and results of prospective cohorts evaluating associations between insulin- or glucose-related intermediate phenotypes and risk of kidney cancer**
